# Multivariate determinants of wearable-measured sleep quality across a large observational cohort: roles of physical activity, gut microbiome, blood analytes, and lifestyle factors

**DOI:** 10.64898/2026.05.27.26354250

**Authors:** Jacob Cavon, Crystal Perez, Nick Quinn-Bohmann, Andrew T. Magis, Sean M. Gibbons

## Abstract

Emerging evidence links the gut microbiome to sleep quality, yet measuring sleep at scale remains challenging. Commercial wearables, such as Fitbit, capture objective sleep and activity data in naturalistic settings. We integrated Fitbit data from a large, deeply-phenotyped cohort with paired lifestyle and health questionnaires. Wearable-derived measures aligned well with self-reported sleep, activity, and happiness. We identified dozens of covariate-adjusted associations between Fitbit-derived sleep features, lifestyle factors, and multi-omic data. Among molecular feature sets, the gut microbiome showed the greatest number of associations with sleep quality: butyrate-producing genera were positively associated with sleep and amplified the benefits of physical activity. *Oscillospira*, in particular, was consistently associated with better sleep. In blood, insulin, omega-3, and cortisol correlated with poorer sleep, whereas lower alcohol intake and mineral supplements correlated with better sleep. These robust, covariate-adjusted findings advance mechanistic understanding of the gut-sleep axis and broader molecular and lifestyle determinants of sleep quality.

## Introduction

Sleep is an essential physiological process that supports brain, cardiovascular, and immune function, as well as metabolism and appetite control ^1^. Disruptions to sleep, including inappropriate timing, irregularity, and aberrant duration have been linked to short- and long-term adverse health consequences ^2^, higher frailty in older individuals ^3,4^, and an increased risk for accidents and mortality ^5,6^. Poor sleep is a critical health issue across the lifespan, with the Centers for Disease Control and Prevention (CDC) estimating that, in the U.S., about one-third of adults and children less than 14-years-old and 75% of high schoolers get less than the recommended amount of sleep for their age categories (https://www.cdc.gov/cdi/indicator-definitions/sleep.html). A recent study using objective monitoring data from over 20 countries found that only 12.9% of individuals achieve both the recommended 7-9 hours of sleep/night and >8,000 steps/day ^7^, and showed a bidirectional coupling between sleep and activity where insufficient sleep duration on the preceding night had a stronger negative effect on next-day activity than the reverse ^7^. Consistent with this, a randomized cross-over trial found that sleep restriction results in less physical activity the following day ^8^. These findings underscore the interdependence between activity and sleep, and the need for targeted sleep interventions that may yield downstream benefits for both physical activity and overall health.

Emerging research has begun to map biomolecular correlates of sleep quality, which could identify mechanistic targets for therapeutic intervention. For instance, a recent metabolomics study in a large U.S. Hispanic/Latino cohort linked blood metabolites, including bile acids, lipids, and amino acids, to multiple self-reported sleep phenotypes, generating one of the first molecular “atlases” of sleep ^9^. Several animal and human observational studies of the gut-sleep axis have linked short-chain fatty acid-producing microbes to sleep quality ^10–14^, and randomized controlled trials have shown probiotics improve sleep quality ^15–18^. Importantly, most of these studies rely on the Pittsburgh Sleep Quality Index (PSQI) or other subjective self-reports, which, although they have the advantage of being easily applied to large cohorts, are prone to recall bias, capture only sleep from the prior month, and measure perception rather than physiology ^19^. Conversely, in studies where objective sleep measurement tools are used, sleep is measured in a clinical setting for only a few days with small sample sizes, which does not capture the heterogeneity of real-world cohorts in an at-home setting.

Indeed, a central challenge in sleep research is objectively measuring sleep at-scale in naturalistic settings. Polysomnography (PSG), the gold-standard, is expensive, low-throughput in terms of number of participants and number of nights that can be recorded at a time, requires technical expertise to administer and analyze, and typically captures sleep data in clinical rather than naturalistic settings ^20^. In contrast, actigraphs, which are wrist-worn devices that monitor movement, offer a key advantage over PSG by enabling continuous and longitudinal at-home sleep assessment for larger sample sizes, but tend to give unreliable estimates for certain sleep parameters ^21^. Commercial wearable devices are emerging as a lower-cost alternative to the research-grade Actiwatch device, and come with additional sensors that can enhance sleep characterization ^22^. Several commercial wearables, including Fitbit, more closely align with PSG measurements than Actiwatch ^23^, overcoming some of the limitations of the Actiwatch. These devices have the advantage of capturing paired longitudinal sleep and activity data from large populations of people in their natural environments.

In this study we calculate monthly averages and standard deviations of a variety of Fitbit sleep and activity metrics for each individual. Fitbit metrics show strong alignment with self-reported sleep, activity, and happiness assessments. As expected, we found a strong association between physical activity and sleep variables. We identify several lifestyle and multi-omic features associated with Fitbit-based monthly average sleep duration, efficiency, disruption, bedtime, and their variabilities, while adjusting for Fitbit-based physical activity metrics, mental illness and sleep disorder diagnoses, medication use, psychological stress, sex, age, body mass index (BMI), genetic ancestry, season, and monthly weekend-weekday ratio variation. In addition to identifying associations independent of physical activity, we examine the interaction effects between physical activity and gut microbe presence/absence on sleep parameters, controlling for the same covariates mentioned above, highlighting potential probiotic targets that may augment the beneficial impacts of activity on sleep quality. Our findings, from a large, real-world cohort with extensive covariate control, provide a foundation for generating mechanistic hypotheses about how lifestyle factors, gut microbiome features, supplements, and host pathways are linked to sleep.

## Results

### Study design and cohort statistics

Study participants were subscribers of the Arivale Scientific Wellness programme and provided consent and research authorization allowing the use of their anonymized, deidentified data in research. Arivale, Inc. (USA) was a consumer scientific wellness company that opened in 2015 and shut down in 2019. The Arivale programme is described in detail in Zubair et al. ^24^. In brief, participants signed up for a comprehensive deep-phenotyping program, which involved collection of comprehensive blood and stool multi-omics, health and lifestyle questionnaires, and wearable data. We conducted a cross-sectional analysis to map associations between lifestyle factors, biological multi-omic data, Fitbit-derived activity measures, and 18 Fitbit-derived sleep features. For each Arivale participant, Fitbit recordings were aggregated as the mean and standard deviation within ±15 days of stool sample collection (Fig. S1). All additional sampled data (e.g. lifestyle and multi-omic data) were merged around the stool sample collection date using time windows chosen to maximize data completeness, while minimizing the time elapsed between measurements (Fig. S1). The subsetted cohort, after data merging, had 1,591 total participants, with some missingness depending on participant data availability. The cohort showed a female bias, with 980 female (62%) and 611 (38%) male participants, with an average age of 47.3±11.7 and an average BMI of 28.0±6.2. Additionally, 54 participants (3.4%) reported taking melatonin, while 74 participants (4.7%) reported taking a sleep drug (Z-drug). There were seven participants who reported taking both melatonin and a Z-drug, resulting in a total of 133 participants (8.4%) who reported taking a Z-drug and/or melatonin. Participants also responded to sleep disorder and mental illness questionnaires. There were 322 participants (20%) who reported any current or previous diagnosis of a sleep disorder, with insomnia being the most prevalent diagnosis. For the mental illness questionnaire, the most prevalent diagnosis was depression, with 529 participants (33%) reporting any current or previous diagnosis of a mental illness.

### Comparing Fitbit sleep and activity metrics against questionnaire data

We internally validated Fitbit sleep and activity features by comparing them against multiple self-reports, including sleep and activity questionnaires, sleep disorder and mental illness questionnaires, and the Oxford Happiness Questionnaire (OHQ). A detailed description of each Fitbit feature, and the Fitbit data processing pipeline, is available in the Fitbit data dictionary and in the Methods section. We fit linear regression models in which individual Fitbit sleep or activity features were treated as continuous dependent variables and sleep and activity questionnaire items were treated as independent variables modeled using an orthogonal polynomial coding scheme (see the Methods section for further details). Sleep disorder and mental illness questionnaires were modeled as outcomes using logistic regression with Fitbit sleep features as continuous predictors. OHQ items were treated as continuous outcomes in linear regression models. All models were adjusted for sex, age, BMI, the first three principal components of genetic ancestry, the number of days between Fitbit recording and questionnaire completion, and the questionnaire vendor. Coefficients and p-values for independent features of interest were grouped by dependent variable, and the Benjamini-Hochberg procedure was applied to each grouping of p-values to control the false discovery rate (FDR), setting the q-value threshold to less than 0.1.

Evaluation of activity questionnaire responses against Fitbit activity features revealed several statistically significant linear relationships (q < 0.1; Fig. 1), where both self-reported vigorous activity duration and moderate activity duration were significantly (q < 0.1) positively associated with all three tested Fitbit activity features (steps, activityCalories, and floors). Fitbit sleep features aligned with the sleep questionnaire (q < 0.1; Fig. 1). Greater self-reported average sleep was positively associated with Fitbit minutes asleep and time in bed. The restful sleep question, which included “yes”, “unsure”, and “no” as possible responses coded in ascending order from 0 to 2, respectively, was associated with 12 out of the 18 total Fitbit sleep metrics. It was negatively associated with average time in bed, minutes asleep, and sleep efficiency (the percentage of time asleep while in bed), and positively associated with average minutes awake, awake duration, restless duration, minutes awake variability, awake duration variability, restless duration variability, time in bed variability, minutes asleep variability, and sleep efficiency variability. Furthermore, self-reported impact of a poor night’s sleep on the ability to concentrate during the day, with possible responses ranging from “not at all” to “an extreme amount of impairment”, was positively associated with sleep efficiency variability and average minutes after wakeup.

**Figure 1:**
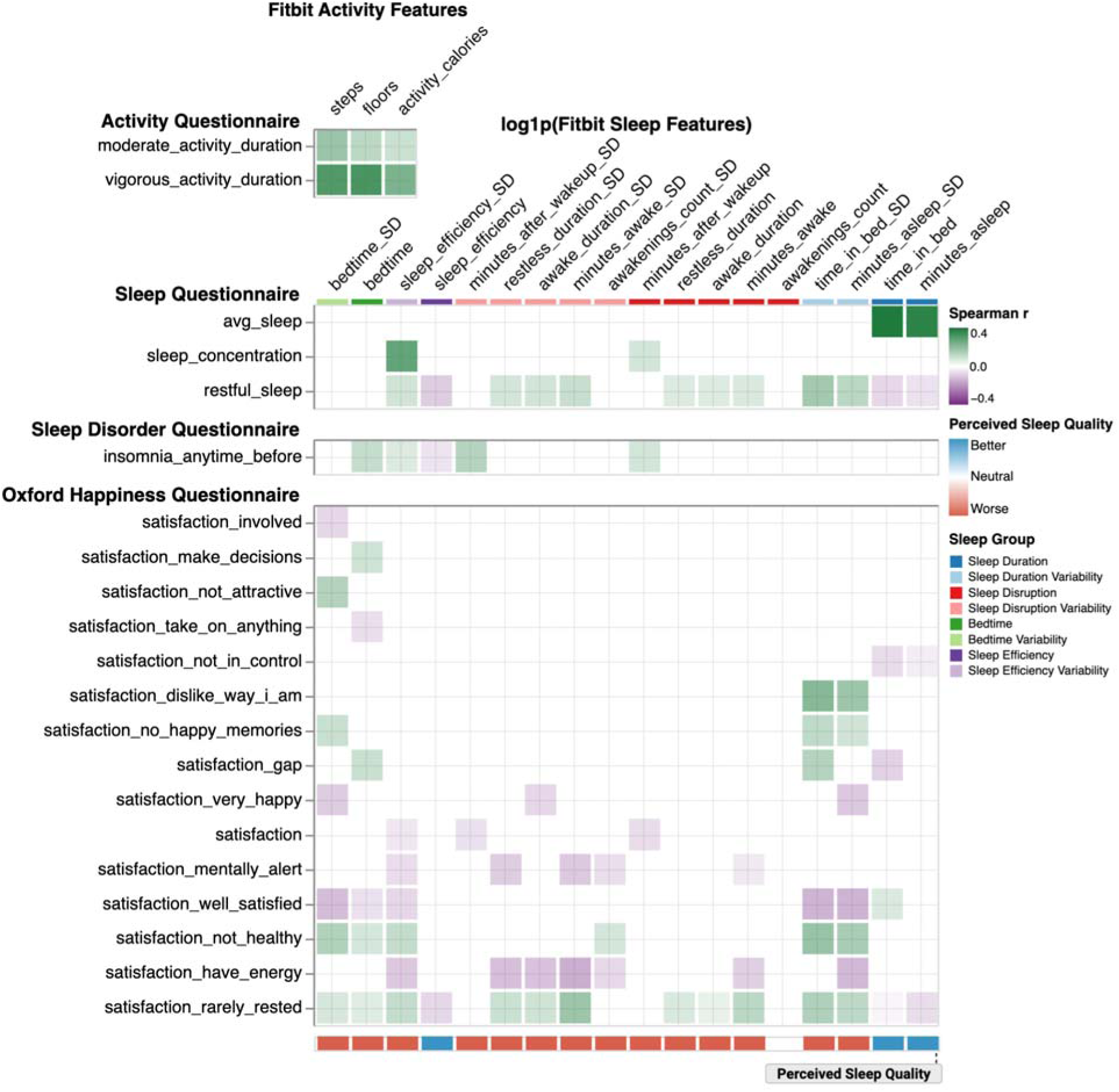
Fitbit features aligned with activity and sleep questionnaires, insomnia diagnosis, and the Oxford Happiness Questionnaire. Heatmap of Spearman correlations of Fitbit-derived sleep and activity features significantly associated (q < 0.1) with activity and sleep questionnaires, insomnia diagnosis, and Oxford Happiness Questionnaire (OHQ) items. Sleep and activity questionnaire items were modeled as ordinal categorical predictors using an orthogonal polynomial coding scheme, where statistical significance was assessed based on the linear term of the ordinal predictor. Insomnia diagnosis was modeled as the outcome in a logistic regression. OHQ items were treated as continuous outcome variables. All models were adjusted for sex, age, BMI, the first three principal components of genetic ancestry, time between Fitbit recording and questionnaire completion, and the questionnaire vendor. Fitbit sleep features were grouped into higher-level sleep domains, indicated by the color bar at the top of the plot. A Perceived Sleep Quality score was computed for each Fitbit sleep feature by summing the direction of significant associations across OHQ items (+1 for positive, −1 for negative, with equal weights for each OHQ question); summed values >0 were labeled “better,” <0 “worse,” and 0 “neutral”, with regard to perceived sleep quality, indicated by the color bar at the bottom of the plot. Sample sizes varied by feature set. Activity Questionnaire: median[IQR] = 1,521[1,520-1,521]; Sleep Questionnaire: 527[177-1,320]; Sleep Disorder Questionnaire: 980 [219-1,591]; OHQ: 936[936-1,499].

Fitbit sleep features were able to weakly discriminate between participants with and without any current or previous diagnosis of insomnia (Fig. 1). Greater Fitbit sleep efficiency variability, average minutes after wakeup and its variability, and later bedtimes were associated with increased odds of any current or previous insomnia diagnosis (q < 0.1). Greater sleep efficiency was associated with decreased odds of insomnia (q < 0.1). For current or previous mental illness diagnoses, greater minutes asleep variability and later bedtimes were associated with increased odds of a depression diagnosis (p < 0.05), but these associations did not survive FDR control (q ≥ 0.1).

Finally, all the Fitbit sleep features, except for awakenings count, were associated with at least one item from the OHQ (q < 0.1; Fig. 1). Responses were on a 1 to 6 Likert-scale where 1 meant “strongly disagree” and 6 meant “strongly agree.” Notably, all except for four of the 18 Fitbit sleep features were associated with the “rarely rested” item (q < 0.1), and in directions logically consistent with the sleep feature. Further, these associations were concordant with the 12 “restful sleep” associations from the Sleep Questionnaire, capturing two additional associations with bedtime and its variability. For instance, minutes asleep was negatively associated with “rarely rested,” meaning more minutes asleep was associated with greater disagreement with that item, echoing the “restful sleep” negative association with minutes asleep. Similarly, sleep efficiency was negatively associated with “rarely rested,” while greater variability in sleep efficiency was positively associated with the same variable. Later bedtimes and larger bedtime variability were also positively associated with “rarely rested.”

We organized the 18 Fitbit sleep features into 8 different sleep variable groups: average sleep duration, average disruption, average bedtime, average efficiency, and the standard deviations of these variables. Furthermore, we used the OHQ to build a perceived sleep quality score, where Fitbit sleep associations with OHQ items that improved the happiness score were coded as “+1,” and those that decreased the happiness score were coded as “-1.” Scored associations were then summed down the columns of Figure 1 for each Fitbit sleep metric, with the final perceived sleep quality score reflecting better, worse, or neutral sleep quality if the sum was greater than, less than, or equal to zero, respectively. In downstream univariate regression analyses, an overall impact score on perceived sleep quality was calculated by summing across significant Fitbit sleep metric associations for each lifestyle and multi-omic predictor, with each sleep metric receiving equal weight (+1, -1, or 0, depending on the OHQ classification).

### Machine learning identifies feature sets that explain the most variance in Fitbit sleep metrics and provide guidance on covariate selection

For all of the following analyses we excluded participants who reported taking any Z-drug and/or melatonin (N = 133). Lifestyle and multi-omics data were organized into 10 different predictor feature sets. Lifestyle feature sets included a daily food frequency questionnaire, self-reported supplements, and a digestion questionnaire. Multi-omics feature sets comprised blood proteomics, untargeted blood metabolomics, blood-based clinical chemistries, and four feature sets derived from 16S stool amplicon sequencing data: gut microbiome alpha diversity metrics, CLR-transformed gut bacterial genus-level abundances, gut bacterial genus presence/absence, and predicted metabolic outputs from participant-specific microbial community-scale metabolic models (MCMMs) constrained at the genus-level using 16S taxonomic profiles and assuming a standard European diet. We first quantified the out-of-sample variance in Fitbit sleep features explained by the predictor feature sets, Fitbit activity and heart rate, and control covariates (sex, age, BMI, genetic ancestry, psychological stress [Perceived Stress Scale; PSS], and history of a sleep disorder or mental illness) alone and in combination with each other (Fig. S2). Random forest models were trained and evaluated using repeated K-Fold cross-validation (four fold splits repeated 10 times: three-folds for training, one-fold for out-of-sample testing; Fig. S2). The total number of samples used to fit and test models varied depending on Fitbit sleep feature outcome, ranging from 497 to 1,469. Missing values in the training and testing folds were imputed with the median of the training folds, with average matrix-wide missingness in testing folds ranging from 0-56% across all feature sets (Table S1). The most missingness was observed in the blood metabolomics feature set. The 2.5-97.5th percentile out-of-sample testing R^2^ distributions show the out-of-sample variance explained in each sleep feature by a feature set (Fig. 2a). Fitbit activity features explained the largest out-of-sample variance in Fitbit sleep outcomes (sleep duration variables, bedtime, and their monthly variabilities) compared to other feature sets. Additionally, all feature sets combined (including Fitbit activity) did not explain more variance in any of the Fitbit sleep features than the Fitbit activity features alone (Fig. 2a). Based on the random forest results, we adjusted for physical activity in all of our downstream univariate regression analyses, to identify activity-independent associations with sleep quality. Furthermore, we ran regression analyses to look for interaction (moderation) effects between activity features and gut microbiome genera (i.e., putatively modifiable components of the phenotype), with regard to their effects on sleep quality. Representative Fitbit activity covariates included steps, floors, and activityCalories, which together cover distance, elevation, and intensity of activity ^25,26^. Downstream models were also adjusted for sex, age, BMI, the first three principal components of genetic ancestry, psychological stress, and history of a sleep disorder or a mental illness. To assess multicollinearity between these control covariates and their relationships with the 18 Fitbit sleep features prior to modeling, we computed a pairwise Spearman correlation matrix (Fig. 2b). Missing values were imputed with medians, and p-values were calculated by transforming the Spearman coefficient into a t-statistic (two-tailed). Un-adjusted p-values less than 0.05 were denoted with an asterisk. On average, we observed weak correlations between control covariates, where 75% of pairings had an absolute Spearman’s rho less than 0.13 (Fig. 2b). The strongest correlations were observed in clusters of sleep outcome variables and activity covariates, measuring similar aspects of sleep or activity. For instance, awake duration was moderately correlated (Spearman rho’s around 0.3) with other sleep disruption variables, such as minutes awake, awakenings count, restless duration, and their monthly variabilities. The Fitbit activity features were moderately correlated with each other (e.g., strongest Spearman’s rho was 0.67 between activityCalories and steps), but weakly correlated with the other control covariates (e.g., strongest correlation was between activityCalories and sex at -0.53 [a negative coefficient indicates reduced activity in females relative to males], followed by floors and BMI at -0.36). The relatively weak correlations between the control covariates justified their joint inclusion in the downstream regression analyses. Control covariates were correlated with several Fitbit sleep outcomes. For instance, being female was correlated with greater minutes asleep and earlier bedtimes, while higher BMI was correlated with worse sleep efficiency, greater minutes asleep, and more sleep disruptions. Older age was correlated with earlier bedtimes, greater variability in bedtime, and worse sleep efficiency. Greater psychological stress was correlated with greater monthly sleep efficiency variability and earlier bedtimes. Reported mental illness was correlated with later bedtimes and more monthly variability in sleep duration variables, while reporting a sleep disorder was correlated with later bedtimes and shorter sleep duration. Interestingly, greater activity (i.e. more steps walked, floors climbed, or active calories burned) was correlated with shorter average sleep duration, but less variability in monthly sleep duration. More steps and floors were correlated with less awake duration and less variability in awake duration.

**Figure 2:**
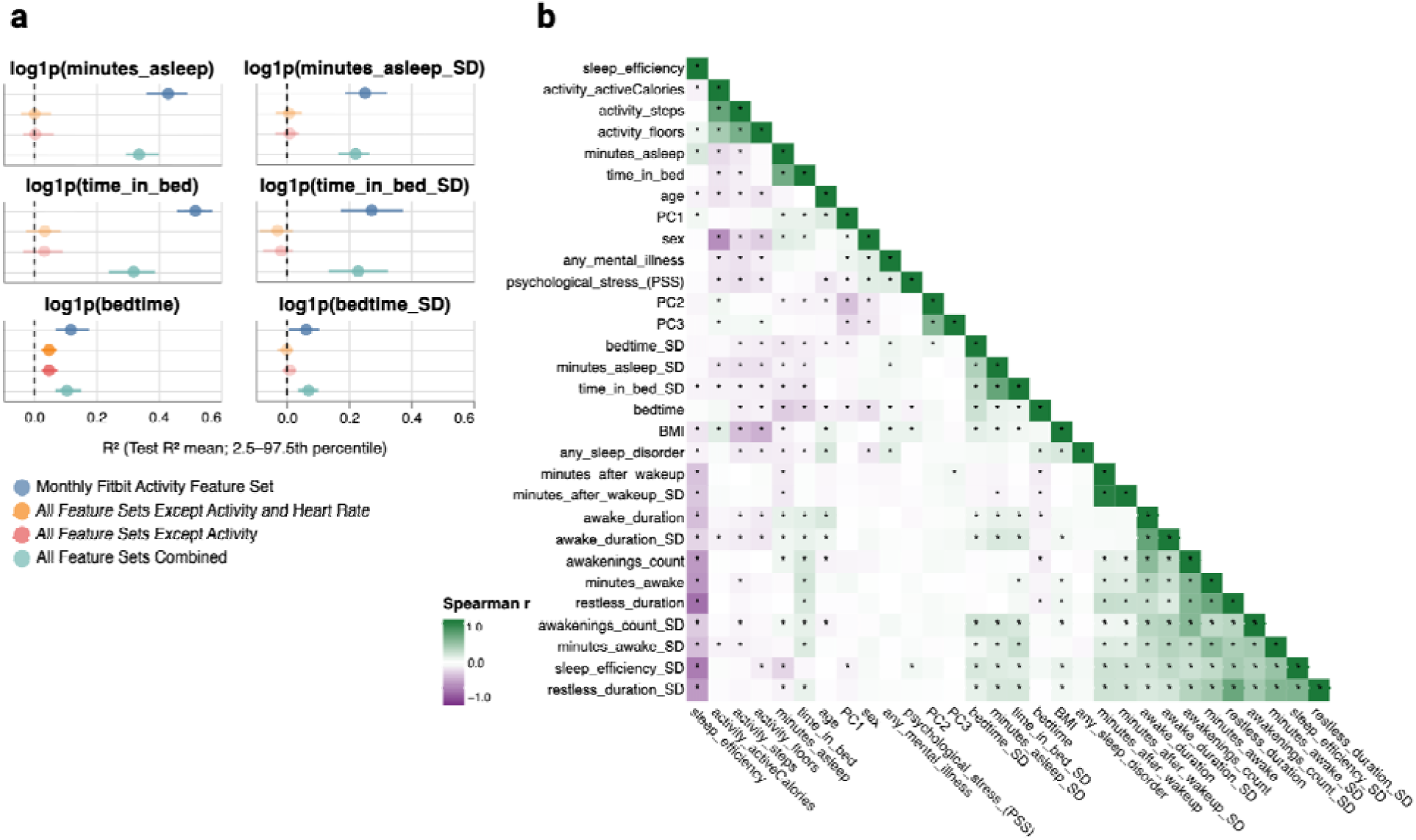
Out-of-sample variance in Fitbit sleep features explained by predictor feature sets, and correlations between Fitbit sleep features and control covariates. (a) Forest plots showing out-of-sample R^2^ distributions across all test folds for random forest models predicting Fitbit sleep outcomes. Colors represent different predictor feature sets used to train random forest models. Each line and point is a different random forest model, where the points represent the out-of-sample R^2^ means, and the lines represent the 2.5th to 97.5th percentile range of each distribution. The full set of tested models (Fig. S2) are not shown, only models whose percentile range did not include 0 for at least one sleep feature are shown. Faded points and lines indicate that the percentile range contains 0. (b) Clustered Spearman correlation matrix between all 18 Fitbit-derived sleep features, three Fitbit-derived activity features (activityCalories, steps, and floors), sex, age, BMI, the first three principal components of genetic ancestry (PC1, PC2, and PC3), history of a sleep disorder or a mental illness, and psychological stress (PSS). Asterisks indicate correlations with p < 0.05. Missing values were imputed as the median of a given variable.

### Diet, digestion, and supplement questionnaire features associated with Fitbit sleep features

To isolate lifestyle and multi-omic variables associated with Fitbit sleep metrics, independent of physical activity, we applied an untargeted discovery-based approach to regress Fitbit sleep features against covariate-adjusted questionnaire-based and multi-omic feature sets (Fig. S3). Many participants had incomplete data, so for each model we only included samples from participants with a complete set of measurements. For each predictor and Fitbit sleep feature outcome, we fit two models: A and B (Fig. S3). Model A estimated the association between the predictor and a given sleep feature while adjusting for a set of baseline covariates: sex, age, BMI, and the first three principal components of genetic ancestry, time elapsed between feature set sample collection and Fitbit recording date, and feature set vendor (when applicable). Model B used the same specification as model A, but additionally adjusted for history of a sleep disorder or a mental illness, psychological stress, and three Fitbit activity features (steps, floors, and activityCalories).

Additionally, to fit a model, we required at least 10 samples per independent variable in the regression formula. For binary predictors we required the number of positive cases to be within 10-90% of the total N used to fit the model, and for ordinal categorical variables, we required each response level to have ≥10 samples. Only predictors with q-value less than 0.1 in both models A and B were considered significant and reported in the figures. See the Methods section for more detail. Model B contained suspected collider variables, which is why we conservatively required significance across both models to retain a feature as a hit. We first examined the daily food frequency questionnaire feature set, making a total of 352 comparisons between 44 different diet questionnaire items and 8 Fitbit sleep features, with sample Ns ranging from 178 to 193 (median[IQR] = 193[193-193]). None of the comparisons with average minutes asleep or time in bed met our minimum N criteria, so diet-sleep duration variable comparisons were not considered in this analysis. The 8 dependent variables tested were monthly average bedtime, awakenings count, sleep efficiency, minutes after wakeup, and the monthly variabilities of these features. We found that more alcoholic drinks consumed per day was associated with a larger bedtime variability (q < 0.1) in both models A and B, which corresponded to worse perceived sleep quality (Fig. 3). All downstream analyses had sufficient N to make comparisons with all 18 Fitbit sleep features. The self-reported supplement feature set contained five different binary variables corresponding to whether a participant’s self-reported supplement-use included mention of a mineral supplement (containing at least one of the keywords: magnesium, calcium, or zinc), multivitamin, omega-3, probiotic, or vitamin D. Predictors were required to meet a minimum of 10% “yes” responses to fit a model, and models had Ns ranging from 497 to 1,469 (median[IQR] = 898[898-1,469]). We found that self-reported use of a mineral supplement was associated with lower awakenings count variability compared to not taking a mineral supplement in both models A and B, corresponding to better perceived sleep quality (q < 0.1; Fig. 3). Several digestion-related questionnaire items associated with different Fitbit sleep features. The questionnaire contained 16 items total, six of which were significantly associated with at least one sleep feature, and the sample sizes of the regressions ranged from 283 to 1,293 (median[IQR] = 797[576-1,072]). Poor appetite was positively associated with later bedtime (Fig. 3). Lack of appetite was also negatively associated with sleep duration features, and greater monthly variability in sleep duration and bedtime, summing together for an overall worse perceived sleep quality. Stress-eating was positively associated with greater average restless duration and its variability. Taking digestion supplements was positively associated with time in bed. Overall, several self-reported digestion features were associated with Fitbit sleep features, where poorer self-reported digestion was generally linked with poorer sleep quality.

**Figure 3:**
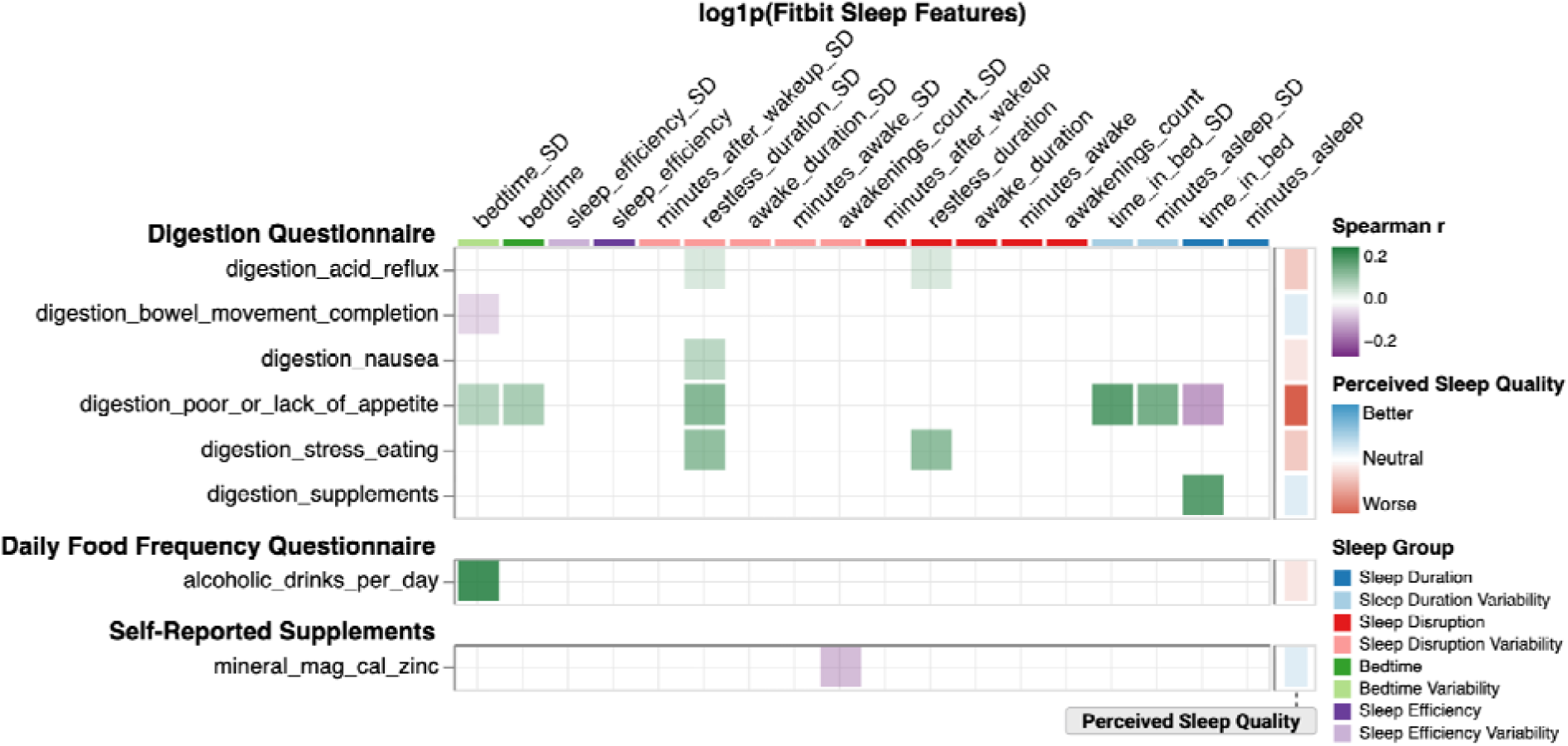
Self-reported digestion, food frequency, and supplement features significantly associated with Fitbit sleep features. Heatmap of Spearman correlations of significant associations (q < 0.1 in models A and B) between digestion, diet, self-reported supplements, and Fitbit sleep features. Fitbit sleep features were organized into higher-level sleep domains, indicated by the color bar at the top of the plot. Perceived Sleep Quality scores were computed for each predictor (y-axis) by summing the equally-weighted direction of its associations across sleep features (+1 or -1 per feature), where sleep feature direction was defined from the Fitbit sleep-OHQ analysis (Fig. 1: higher OHQ = “better,” lower OHQ = “worse”), displayed as the vertical color bar on the right of the plot. Sample sizes varied by feature set. Digestion Questionnaire: median[IQR] = 797[576-1,072]; Daily Food Frequency Questionnaire 193[193-193]; Self-Reported Supplements 898[898-1,469].

### Blood plasma-based proteomics

A total of 4,948 different blood proteomics to Fitbit sleep feature comparisons were made by testing 276 unique proteomics features against the 18 Fitbit sleep features. The set of fitted regression models had Ns ranging from 215 to 873 (median[IQR] = 584[583-872]). We found no associations between any of the proteomics features with any of the Fitbit sleep features that retained significance across both models A and B.

### Blood plasma-based untargeted metabolomics

For untargeted blood metabolomics, we made a total of 20,788 comparisons between metabolite and Fitbit sleep features. The fitted regressions had Ns ranging from 159 to 720 (median[IQR] = 470[264-684]). Of the 1,191 unique metabolites tested, there were three metabolites that were significantly associated with a sleep feature (q < 0.1) in both models A and B (Fig. 4). Blood cortisol was the only blood metabolite associated with bedtime, showing a positive association with later bedtime (Fig. 4). An unidentified blood metabolite (X - 12816) that has been associated with coffee intake ^27^ was negatively associated with minutes after wakeup and its variability. Finally, docosahexaenoylcarnitine (C22:6) was positively associated with restless duration variability and minutes awake variability (q < 0.1), which translated to worse perceived sleep quality.

**Figure 4:**
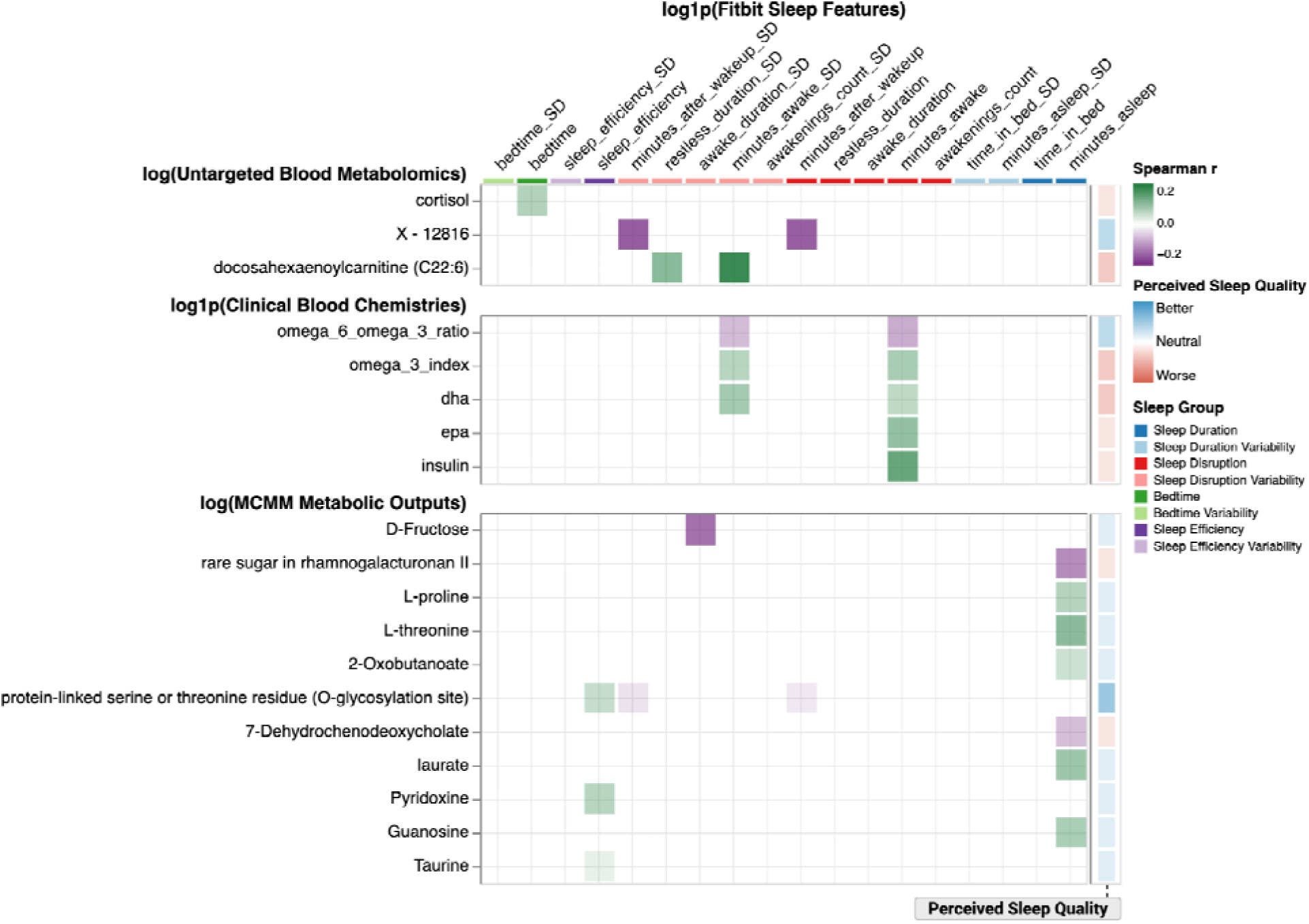
Untargeted blood metabolomics, clinical blood chemistries, and MCMM-**predicted gut metabolic outputs significantly associated with Fitbit sleep features.** Heatmap of Spearman correlations of significant associations (q < 0.1 in models A and B) between untargeted blood metabolomics, clinical blood chemistries, MCMM-predicted metabolic outputs from the gut under a standard European diet, and Fitbit sleep features. Fitbit sleep features were organized into higher-level sleep domains, indicated by the color bar at the top of the plot. Perceived Sleep Quality scores were computed for each predictor (y-axis) by summing the equally-weighted direction of its associations across sleep features (+1 or -1 per feature), where sleep feature direction was defined from the Fitbit sleep-OHQ analysis (Fig. 1: higher OHQ = “better,” lower OHQ = “worse”), displayed as the vertical color bar on the right of the plot. Sample sizes varied by feature set. Untargeted Blood Metabolomics: median[IQR] = 470[264-684]; Clinical Blood Chemistries 734[408-1,127]; MCMM Metabolic Outputs 898[898-1,466].

### Blood-based clinical chemistries

For clinical chemistries, we tested 104 unique features against all 18 Fitbit sleep features for a total of 1,770 comparisons. The fitted regressions had Ns ranging from 163 to 1,147 (median[IQR] = 734[408-1,127]). Consistent with the DHA-carnitine conjugate finding from the metabolomics feature set (docosahexaenoylcarnitine), measures of blood omega-3 fatty acids were positively associated with sleep disruption and its variability (Fig. 4). For instance, the omega-3 index, DHA, and EPA were all positively associated with average minutes awake, corresponding to worse perceived sleep quality, while the omega-6 to omega-3 ratio was negatively associated with average minutes awake (q < 0.1). Omega-3 index and DHA were positively associated with the standard deviation of minutes awake, while the omega-6 to omega-3 ratio was negatively associated with the standard deviation of minutes awake (q < 0.1). Higher fasting insulin was associated with greater minutes awake (q < 0.1).

### Stool 16S amplicon sequencing

Using each participant’s stool 16S amplicon sequencing data we constructed genus-level microbial community-scale metabolic models (MCMMs) using MICOM (v0.37.0) ^28^. Model exchanges were constrained using an assumed standard European diet, and growth simulated with cooperative tradeoff flux balance analysis. Metabolic production rates, predicted by MICOM, represent the community-wide export flux of a given metabolite into the luminal compartment (https://micom-dev.github.io/micom/autoapi/micom/measures/index.html). We compared a total of 201 predicted metabolic exports with all 18 Fitbit sleep features, for a total of 3,534 unique metabolic output-sleep feature comparisons. Regression models had Ns ranging from 171 to 1,469 (median[IQR] = 898[497-1,466]). We identified 11 metabolite export flux associations that were significant in both models A and B (Fig. 4). D-Fructose was negatively associated with awake duration variability translating to better perceived sleep quality, while a rare sugar from rhamnogalacturonan II was negatively associated with minutes asleep, which corresponded to worse perceived sleep quality. Conversely, amino acids L-threonine and L-proline, as well as the L-threonine degradation product 2-Oxobutanoate, were positively associated with minutes asleep and better perceived sleep quality. Complementary to this, protein-linked serine and threonine production fluxes were associated with better sleep efficiency and decreased minutes after wakeup and its variability. Laurate and guanosine production fluxes were both positively associated with minutes asleep. Pyridoxine (vitamin B6) and taurine production fluxes were both positively associated with sleep efficiency and therefore better perceived sleep quality, while the bile acid 7-dehydrochenodeoxycholate was negatively associated with minutes asleep, corresponding to worse perceived sleep quality. We tested three alpha diversity metrics: the number of unique observed features (i.e. taxon richness), the Berger Parker index (i.e., an evenness metric), and the Shannon index (i.e., a metric that integrates both richness and evenness). All three alpha diversity metrics were significantly associated with Fitbit sleep features (q < 0.1) in both models A and B (Fig. 5), with Ns ranging from 497 to 1,469 (median[IQR] = 898[898-1,469]) (Fig. 5). The number of observed features and the Shannon index were negatively associated with awake duration, awake duration variability, awakenings count, time in bed, and minutes asleep, translating to an overall neutral impact on perceived sleep quality. The Berger Parker index was positively associated with these same Fitbit sleep features. Next, we identified associations between CLR-transformed genus-level gut bacterial abundance features and Fitbit sleep features (Fig. 5). We made a total of 2,744 comparisons looking at 177 different microbial genera against all 18 sleep features. The regression models had Ns ranging from 170 to 1,469 (median[IQR] = 509[325-875]). There were 15 unique microbial genera, mostly from the Ruminococacceae and Lachnospiraceae families, that were associated (q < 0.1) with a sleep feature in both models A and B. (Fig. 5). The majority of these associations were with sleep duration variables and tended to be positively related to sleep quality. All genera, except for *Clostridium_sensu_stricto_1* and *Lachnospiraceae_UC5_1_2E3*, were positively associated with sleep duration. *Clostridium_sensu_stricto_1* was negatively associated with sleep duration, corresponding to worse perceived sleep quality, while *Lachnoclostridium* was the only genus with associations across different sleep variable groups, resulting in a neutral impact on perceived sleep quality. Specifically, *Lachnoclostridium* was positively associated with both sleep duration features, awake duration, and awake duration variability. *Lachnospiraceae_UC5_1_2E3* was the only other genera positively associated with awake duration variability, corresponding to worse perceived sleep quality. Besides genera from the Ruminococacceae and Lachnospiraceae families, *Bacteroides, Bilophila, Holdemania,* and *Parabacteroides*were positively associated with minutes asleep.

**Figure 5:**
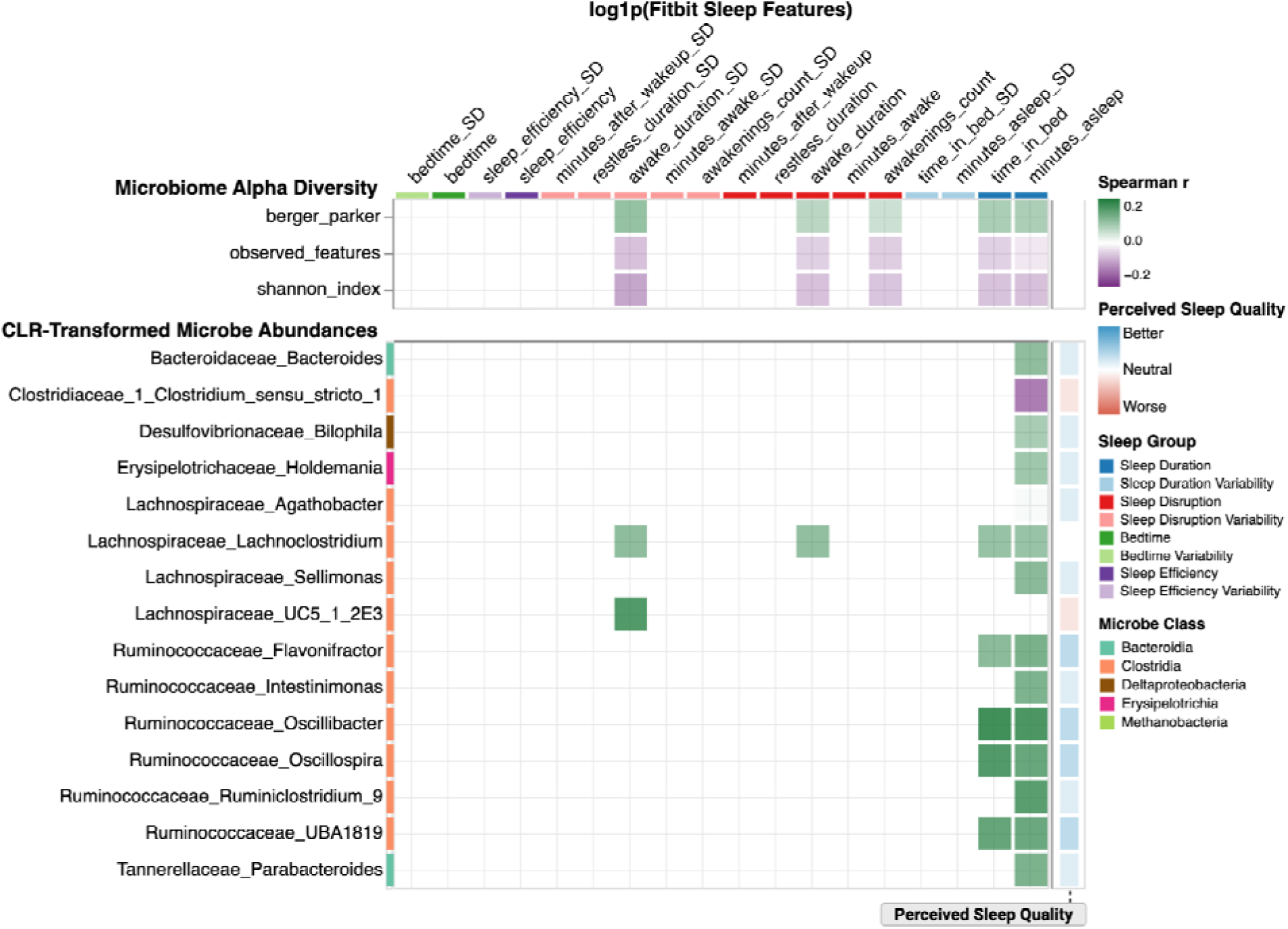
Gut microbiome alpha diversity and CLR-transformed microbe abundance features significantly associated with Fitbit sleep features. Heatmap of Spearman correlations of significant associations (q < 0.1 in models A and B) between gut microbiome alpha diversities, CLR-transformed microbe abundances, and Fitbit sleep features. Fitbit sleep features were organized into higher-level sleep domains, indicated by the color bar at the top of the plot. Microbe features are labeled with their taxonomic class assignment on the left color bar. Perceived Sleep Quality scores were computed for each predictor (y-axis) by summing the equally-weighted direction of its associations across sleep features (+1 or -1 per feature), where sleep feature direction was defined from the Fitbit sleep-OHQ analysis (Fig. 1: higher OHQ = “better,” lower OHQ = “worse”), displayed as the vertical color bar on the right of the plot. Sample sizes varied by feature set. Microbiome Alpha Diversity: median[IQR] = 898[898-1,469]; CLR-Transformed Microbe Abundances 509[325-875].

Genera from the Ruminococacceae and Lachnospiraceae families also predominated the microbe presence/absence feature set, in terms of significant associations with sleep variables (q < 0.1 in models A and B; Fig. 6). We made a total of 2,714 comparisons looking at 154 microbe presence/absence features against all 18 sleep features, and regression models had Ns ranging from 497 to 1,469 (median[IQR] = 898[898-1,469]). Most features were associated with sleep disruption and tended to be negative associations. Only two genera, *Oscillospira* and *Lachnospiraceae_UC5_1_2E3*, were significantly associated with a sleep feature in both the CLR-transformed and presence/absence feature sets. In general, presence of a taxon was primarily associated with less awake duration, except for *Lachnospiraceae_UC5_1_2E3* and *Erysipoletrichaeceae_Canditus_Stoquefichus*, which were associated with longer awake duration, corresponding to worse perceived sleep quality. Several other genera not belonging to the Ruminococacceae and Lachnospiraceae families were negatively associated with awake duration: *Christensenellaceae_R_7_group*, *Clostridiales_vadinBB60_group*, *Methanobrevibacter*, *Romboutsia,* and *Prevotella_7*, translating to better perceived sleep quality. Presence of an unclassified Clostridia taxon and *Clostridiales_Family_XIII_AD3011_group* were both positively associated with sleep efficiency, while presence of *Coprococcus_2* was negatively associated with minutes asleep variability. There were four taxa with associations across two different sleep groups. *Christensenellaceae_R_7_group* was positively associated with sleep efficiency while being negatively associated with awake duration. *Coprococcus_3* was negatively associated with awake duration and its variability. Finally, *Ruminococcaceae_UCG_010* was negatively associated with minutes asleep variability and awake duration, while *Ruminococcaceae_UCG_014* was negatively associated with awake duration and its variability. Overall, multi-omic analysis revealed that different ‘omic feature sets tended to be related to distinct Fitbit sleep variable groups, impacting different aspects of perceived sleep quality (Fig. 7). Additionally, within feature sets, individual associations with Fitbit sleep features tended to be biased towards one direction (e.g., the microbe presence/absence feature set primarily consisted of negative associations).

**Figure 6:**
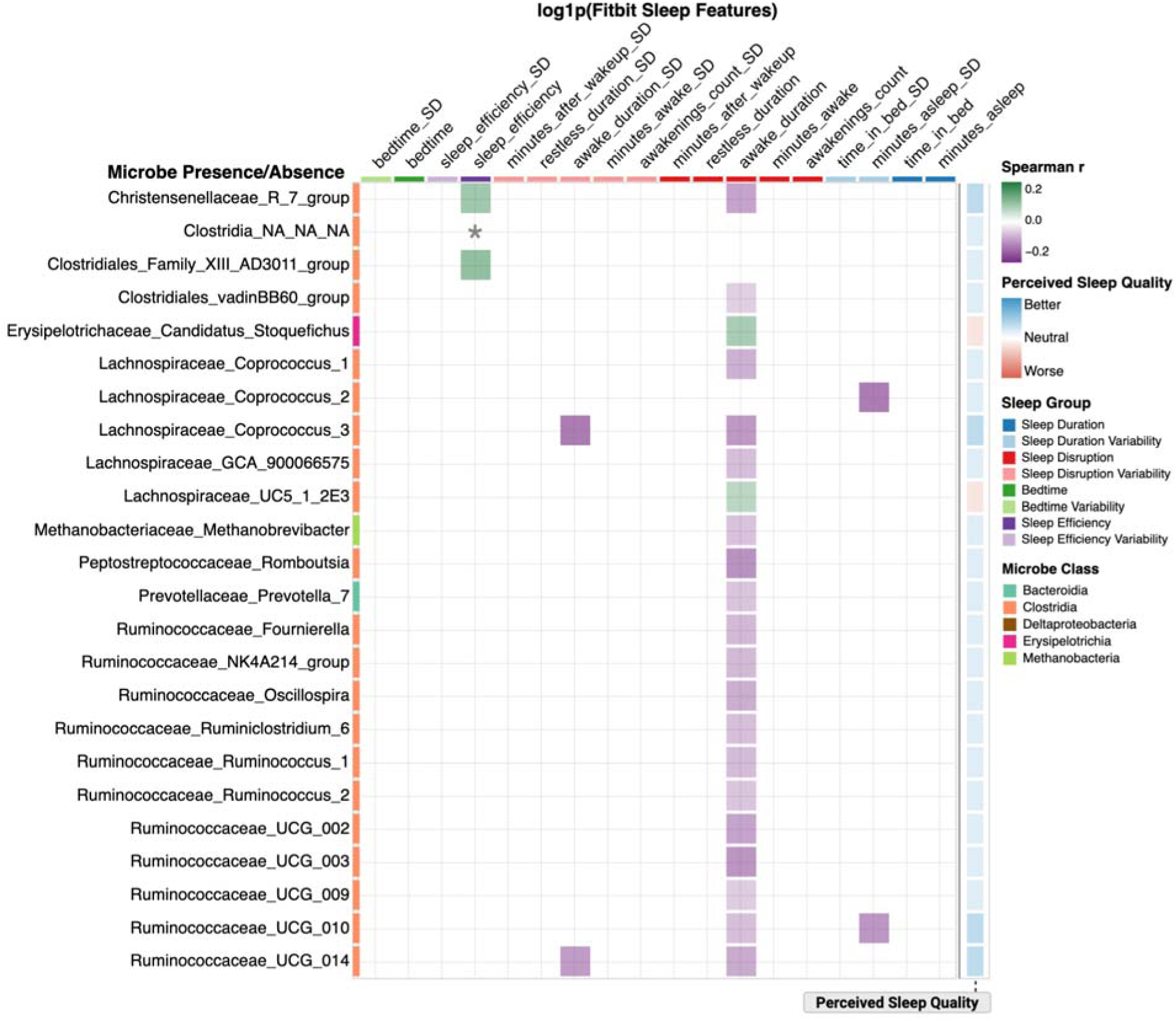
Gut microbe presence/absence features significantly associated with Fitbit sleep features. Heatmap of Spearman correlations of significant associations (q < 0.1 in models A and B) between gut microbe presence/absence and Fitbit sleep features. “*” Spearman r = 0.0014. Fitbit sleep features were organized into higher-level sleep domains, indicated by the color bar at the top of the plot. Microbe features are colored by their taxonomic class assignment on the left color bar. Perceived Sleep Quality scores were computed for each predictor (y-axis) by summing the equally-weighted direction of its associations across sleep features (+1 or -1 per feature), where sleep feature direction was defined from the Fitbit sleep-OHQ analysis (Fig. 1: higher OHQ = “better,” lower OHQ = “worse”), displayed as the vertical color bar on the right of the plot. Sample sizes varied: median[IQR] = 898[898-1,469].

**Figure 7:**
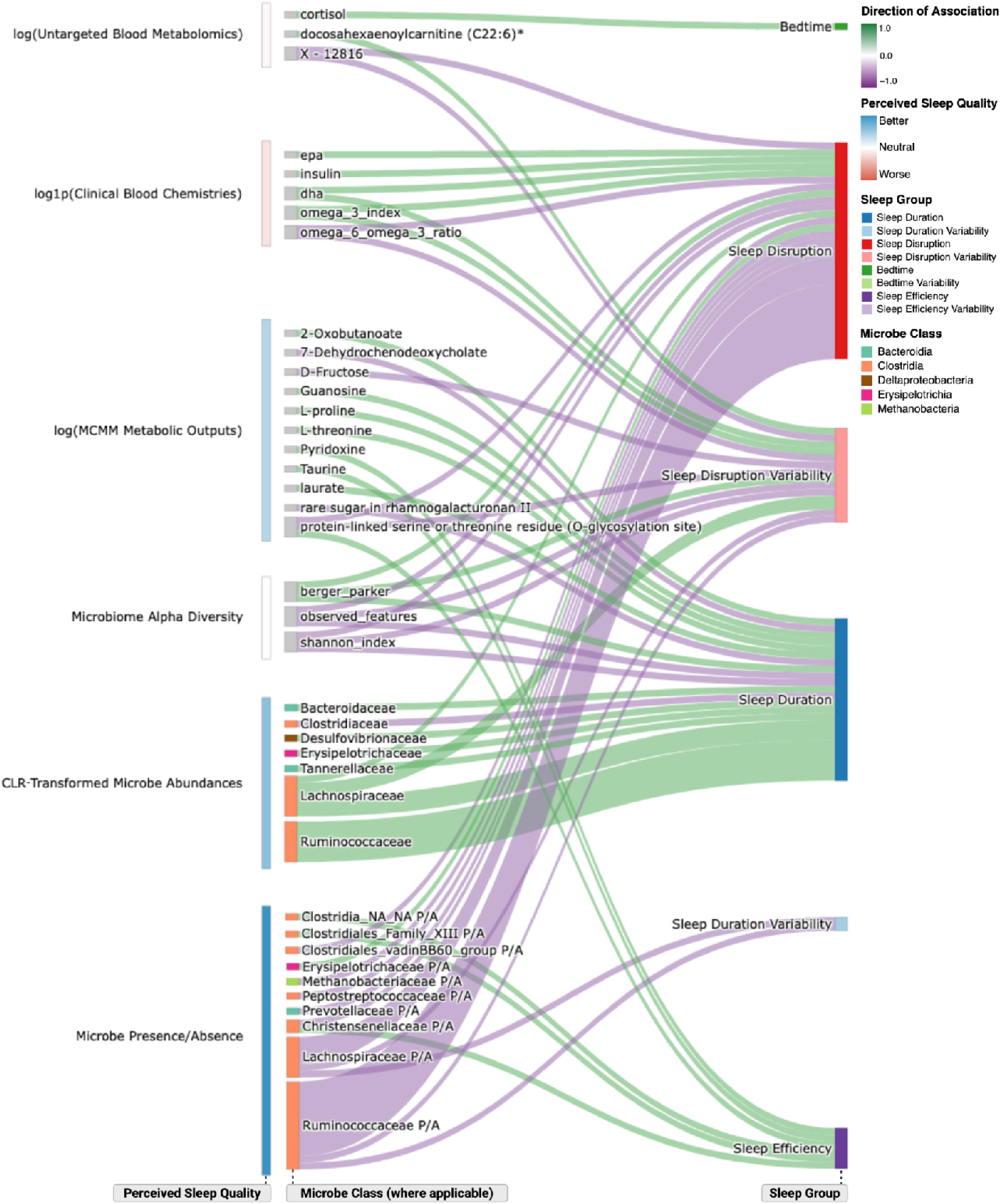
Summary Sankey plot of gut microbiome and blood features significantly associated with Fitbit sleep features. Overview of biological multi-omic features significantly associated with Fitbit sleep features (all results from main Figures 4-6), where Fitbit sleep features have been collapsed into larger sleep groups. Sleep group and predictor feature nodes follow the same coloring convention as in the main Figures 4-6, along with edge color, which indicates direction of association. For microbial features, results were collapsed and summarized at family-level taxonomy, where the edge width of these nodes is proportional to the number of significant microbial genera within a given family. Color bars on the left of the plot represent the total summed Perceived Sleep Quality score of each feature set.

### Gut microbes modify the effect of Fitbit step count and activityCalories on Fitbit sleep features

In addition to the main effects presented above, we looked at interaction effects between the three Fitbit activity features (steps, floors, and activityCalories) with 154 different presence/absence microbes for all 18 sleep features (Fig. S4), fitting a total of 8,142 regressions. Sample sizes ranged from 497 to 1,469 (median[IQR] = 898[898-1,469]). There were 24 total significant activity × microbe interaction terms (q < 0.1) from models containing the steps or activityCalories features as predictors, and awake duration, awake duration variability, bedtime variability, sleep efficiency variability, or minutes asleep variability as outcomes, after adjusting for our full set of covariates (Fig. 8). In models with bedtime variability as the outcome, presence of *Clostridiales_Family_XIII_AD3011_group, Oscillospira, Pseudoflavonifractor, Shuttleworthia,* and *Coprobacter* reversed the steps slope from positive to negative, corresponding to steps improving perceived sleep quality in participants with the microbe present compared to those without the microbe (Fig. 8). In awake duration and awake duration variability models, presence of *Ruminococcaceae_UCG_005* and *Ruminococcus_1* pushed the steps slope from negative to near zero, translating to worse perceived sleep quality with more steps in participants with those taxa present. Presence of *Coprococcus_3, Lachnospiraceae_ND3007_group, Ruminococcaceae_UCG_003,* a genus from the Muribaculaceae family, *Ruminococcaceae_NK4A214_group, Clostridiaceae_1_Clostridium_sensu_stricto_1, Parasutterella, Ruminiclostridium_6, Peptococcus,* and an unclassified genus from the Clostridiales order shifted the steps slope from negative to positive (or near zero) in the awake duration models, meaning that in the presence of those taxa, the effect of steps on sleep went from better to worse perceived sleep quality, relative to participants that lacked these genera absent. In contrast, the presence of *Hungatella* and *Eggerthella* shifted the relationship between steps and awake duration variability from positive to negative, corresponding to better perceived sleep quality with more steps in participants where those taxa were present. The Peptococcaceae family modified the effect of activityCalories on sleep efficiency variability from positive to negative. Finally, the presence of *Eggerthellaceae_DNF00809* and *Escherichia_Shigella* modified the effect of activityCalories on minutes asleep variability, with the former shifting the effect to less negative (worse perceived sleep quality), and the latter making the effect more negative (better perceived sleep quality).

**Figure 8:**
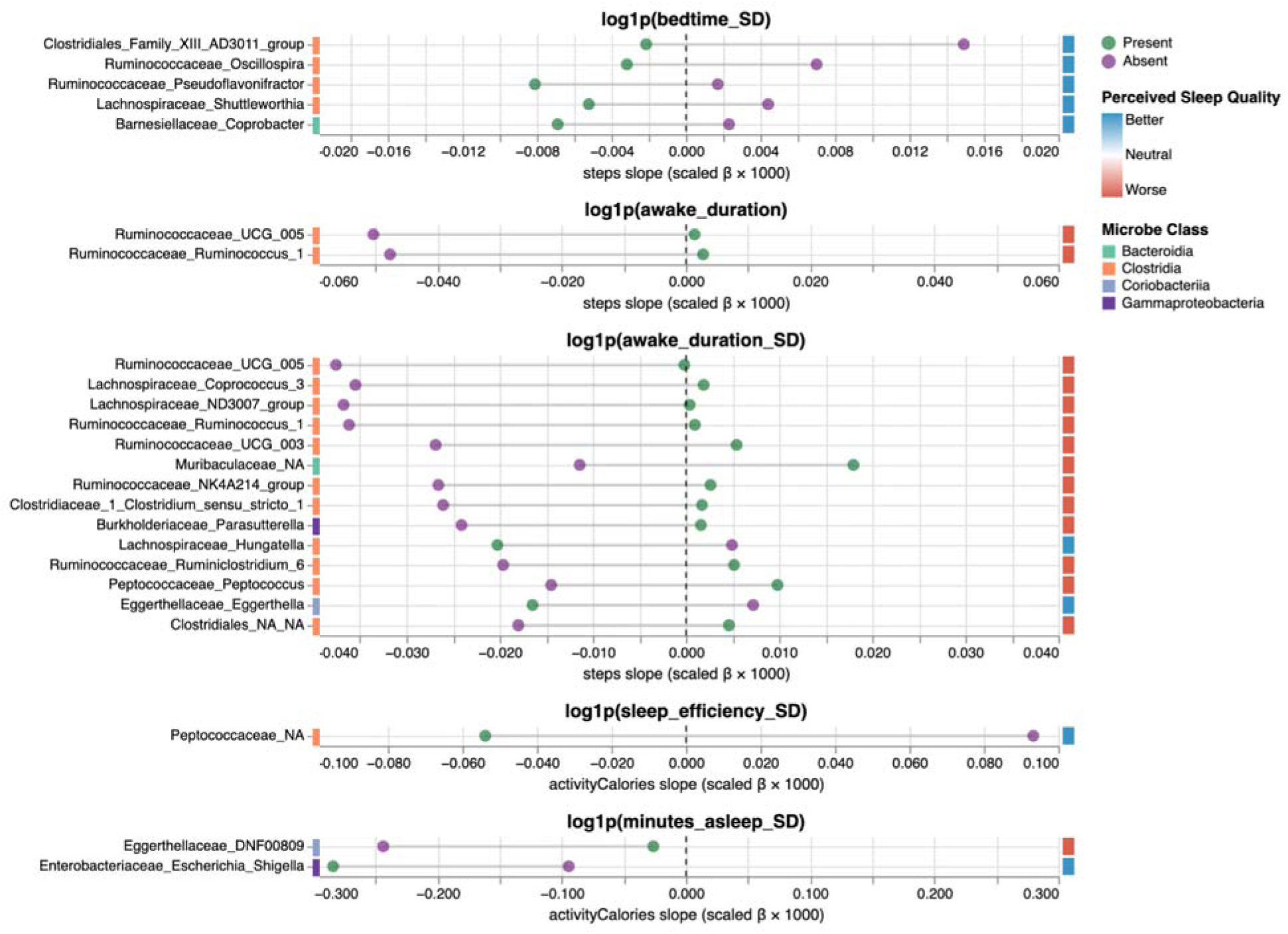
Fitbit Activity × Microbe Presence/Absence interaction effects on Fitbit sleep features. Interaction plots show the estimated Fitbit activity coefficient conditional on the absence or presence of each microbe. Points are colored by microbe presence/absence, and represent the estimated activity coefficients under each condition. Connecting lines correspond to the magnitude of the interaction effect coefficient. The y-axis was organized in descending order from largest to smallest interaction effects from top to bottom within each subplot. The taxonomic class of each taxon on the y-axis was indicated by the color bar on the left. The right-most color bar denotes whether the presence of a microbe improves or worsens Fitbit activity’s effect on sleep, based on the perceived sleep quality score derived from the OHQ, relative to microbe absence. All coefficients are scaled by 1000 for visualization. Only interaction effects with q < 0.1 are shown. All models were adjusted for sex, age, BMI, the first three principal components of genetic ancestry, microbiome vendor, time elapsed between feature set sample collection and Fitbit recording date, presence of sleep disorder or mental illness and the corresponding questionnaire vendor, and psychological stress. Sample sizes varied: median [IQR] = 898[898-1,469].

## Discussion

Here we demonstrated the utility of Fitbit trackers as a means of reliably capturing monthly averages and standard deviations of sleep and activity metrics in the large, deeply-phenotyped Arivale cohort. Fitbit metrics recapitulated epidemiological findings, such as longer sleep durations in women compared to men ^29^, decreased sleep efficiency with age ^30^, and decreased sleep duration with more steps ^31^. Further, a previous paper looked at an earlier subset of Arivale participants, applying a different analytical framework, and found that sleep duration was negatively correlated with BMI, mirroring findings from the current analysis ^32^. Consistent with prior work showing concordance between subjective and objective sleep measurement tools ^33^, Fitbit-derived features aligned with sleep and physical activity assessments, the Oxford Happiness Questionnaire, and insomnia diagnosis, capturing several aspects of sleep quality that are distinct, yet complementary ^21^. In particular, monthly Fitbit standard deviation metrics captured longitudinal information about sleep regularity not available in the self-reports that corresponded to worse perceived sleep quality. These metrics are clinically relevant, as a recent study found that objectively-measured sleep regularity was a stronger predictor of all-cause mortality than duration ^34^, and emerging evidence has shown a connection between objectively measured sleep irregularity and adverse health outcomes ^35,36^. By extending beyond conventional averages to include intra-individual standard deviations of sleep duration, disruption, efficiency, and bedtime, our analytical framework revealed dozens of novel behavioral and biological associations with sleep variability.

Activity metrics emerged as the strongest global predictor of sleep. Fitbit activity features explained the most variance in Fitbit sleep duration, bedtime, and their monthly variabilities (Fig. 2a), consistent with a large body of literature showing a strong coupling between sleep and physical activity ^37^. Notably, we found that other feature sets combined with Fitbit activity features did not explain more variance in sleep parameters than the Fitbit activity features alone, suggesting strong interdependence between activity, lifestyle factors, demographics, biological multi-omics, and sleep. Thus, in order to regress non-activity feature sets against sleep variables, it was critical to adjust for activity (and other relevant covariates) to identify robust, activity-independent associations. Starting with our diet, lifestyle, and supplement analysis, we found that higher daily alcohol consumption was associated with larger bedtime variability, consistent with literature showing that treatment of alcohol use disorder results in less sleep onset variability ^38^. In contrast, a study of college students found that greater alcohol consumption is associated with later bedtimes, but not with bedtime variability ^39^. These contradictory results may reflect different effects across the lifespan. In our study, we controlled for age as a covariate, suggesting that alcohol-induced sleep variability is independent of adult life-stage. Furthermore, taking a supplement containing any combination of zinc, calcium, or magnesium was associated with less variability in night time awakenings corresponding to better perceived sleep quality, consistent with randomized controlled trial results finding improved sleep efficiency and PSQI scores with treatment of zinc-rich food or supplements containing either zinc or magnesium ^40–42^.

Our findings from the untargeted blood plasma metabolomics and blood-based clinical chemistry analyses recapitulated findings in the literature. For instance, blood cortisol has been shown to be positively associated with later sleep midpoints, consistent with our finding that cortisol was positively associated with later bedtimes ^43^. Blood insulin levels were positively associated with awake duration, which is consistent with literature showing insulin insensitivity in response to sleep deprivation ^44^. Our study also found that omega-3 fatty acids were positively associated with sleep disruption and its variability, whereas the omega 6:3 ratio showed the opposite associations. This is contradictory to prior studies showing that consumption of foods high in omega-3, such as fish or supplements, improves sleep efficiency, which is a metric that combines sleep duration and sleep disruption parameters (total time asleep divided by total time spent in bed), although some participants with improved efficiency reported feeling less energetic ^45,46^. A recent meta-analysis found that omega-3 supplementation had consistent beneficial effects on infant sleep, but that effects on adult sleep quality were inconclusive, suggesting variable effects across individuals and across the lifespan ^47^. There may be a non-linear relationship between omega-3 consumption and sleep duration, with a protective effect against both very short and very long sleep duration ^48^. Overly long sleep durations, >8 hours, have also been associated with increased morbidity and mortality ^49^. More work is needed to better understand how omega-3 fatty acids impact sleep quality across different contexts. We observed no consistent association between overall perceived sleep quality and covariate-adjusted gut microbiome alpha diversity. A prior meta-analysis of 20 sleep deprivation studies in both humans and non-human animals found that sleep deprivation was associated with lower gut microbiome Shannon alpha diversity ^50^, and a large observational actigraphy study found that higher Shannon diversity and richness were associated with less sleep duration variability and less wake after sleep onset (WASO), although similar to our results, the Shannon diversity-WASO associations did not survive adjustment for BMI and exercise levels ^51^. Differences in findings might be attributed to different covariate adjustments, population characteristics, and statistical analyses. Paradoxically, we found that sleep duration, disruption, and disruption variability showed similar association patterns with alpha-diversity. One explanation for this result is that longer sleep duration provides more opportunity for disruption (Fig. 2b). Furthermore, average sleep disruption and the standard deviation of disruption positively scale with one another, which is a common property of many statistical distributions.

For MCMM outputs, we found that L-threonine and taurine were positively associated with sleep duration and efficiency, respectively, controlling for the full set of covariates. Both of these molecules have demonstrated somnogenic effects in fruit flies ^52,53^. We further found that 7-dehydrochenodeoxycholate, a bile acid, was negatively associated with minutes asleep, translating to worse perceived sleep quality, consistent with a study in patients with chronic liver disease, which found that increased blood serum bile acid concentration was associated with shorter total sleep time and more fragmented sleep ^54^. Finally, while we did not see any significant association with MCMM-predicted butyrate on a standard European diet, we do see consistent associations between butyate producing genera and sleep ^55,56^ (Fig. 5-6). MCMM predictions were limited by the use of a standard diet for all participants, which did not accurately reflect individual variation in dietary intake of butyrogenic substrates. Butyrate has been shown to increase non-REM sleep in several non-human animals ^57^, correct aberrant sleep architecture in mouse models of Parkinson’s disease ^58^ and insomnia ^10^, and improve PSQI scores in people with ulcerative colitis ^59^.

As mentioned above, genera from Lachnospiraceae and Rumminococcaceae families, which consist largely of butyrate producers ^55,56^, comprised most of the gut microbiome hits that showed covariate-adjusted associations with sleep. This is consistent with a recent meta-analysis that found depletion of Lachnospiraceae and Ruminococcaceae was associated with sleep deprivation ^50^, and another review article showing that perturbations of SCFA-producing microbes have generally been associated with poor perceived sleep quality ^60^. We found that the genus *Holdemania* was positively associated with minutes asleep, consistent with a cross-sectional study showing *Holdemania* was negatively associated with the number of nighttime awakenings ^12^. A probiotic randomized-controlled trial (RCT) in older adults with mild cognitive impairment showed enrichment of Erysipelotrichaceae and *Coprococcus* in the treatment group compared to controls, alongside improved PSQI ^15^, consistent with our *Holdemania* (a genus within the Erysipelotrichaceae family) and *Coprococcus* 1-3 findings. We found increasing abundance of *Bacteroides* was associated with more minutes asleep, translating to better perceived sleep quality, which is consistent with a prior prebiotic intervention trial in perimenopausal women that showed an enrichment of *Bacteroides* within the treatment group alongside improved PSQI scores ^61^. However, *Bacteroides* has been shown to be enriched in insomnia patients ^62^ and following sleep restriction ^63^. Similarly, there are mixed findings on *Prevotella*’s role in sleep. We found that *Prevotella* presence was associated with decreased awake duration, which is consistent with enrichment of the Prevetellaceae family in the treatment group of a probiotic trial, compared to controls, alongside improved PSQI scores ^15^, but contradicts prebiotic intervention ^61^ and cross-sectional ^11^ findings showing depletion of *Prevotella* with improved PSQI scores. There is speculation that an increased abundance of *Prevotella* may compensate against poor sleep through increased succinate production ^11^, although inconsistent findings could also be attributed to species-level functional differences, gut ecological context, or variation in diet. Interestingly, several of our microbial taxa associations showed directionally concordant overlap with taxa previously associated with depressive symptoms in the Rotterdam Study (RS) and HELIUS cohorts ^64^. For instance, presence of *Coprococcus_1, Lachnospiraceae_UCG_001*, and *Ruminococcaceae_UCG_003* were associated with decreased sleep duration variability and therefore better perceived sleep quality, consistent with their negative associations with depressive symptoms in the RS and HELIUS cohorts. Similarly, *Christensenellaceae_R7_group* was associated with better sleep efficiency and decreased sleep disruption, and *Parabacteroides* was associated with longer sleep duration, also consistent with these other cohorts. Finally, we found that increasing abundance of *Lachnoclostridium* was associated with greater sleep disruption, sleep disruption variability, and sleep duration, resulting in an overall neutral impact on perceived sleep quality. This taxon has been positively associated with depressive symptoms ^64^ and insomnia risk ^65^, but it has also been associated with better sleep and cognitive performance in elderly adults ^66^, suggesting possible disease- and age-dependent effects. Beyond the gut bacterial main effects observed in our analysis, we identified several Fitbit activity × microbe presence/absence interaction effects. Specifically, the presence/absence of gut microbial taxa modified the effect of steps and the number of calories burned during physical activity on sleep awake duration, awake duration variability, minutes asleep variability, sleep efficiency variability, and bedtime variability. Although recent studies have highlighted activity effect modification on sleep by sex, activity timing, and activity intensity ^37,67^, none have considered molecular phenotypic interaction effects. Our study introduces the gut microbiome as an important moderator of activity and its effects on sleep. Significant activity × microbe interaction effects primarily consisted of microbes belonging to the Lachnospiraceae and Ruminococcaceae families, although unlike the microbial abundance and presence/absence main effects results, genera within these families differentially moderated activity’s effect on sleep leading to both better and worse sleep quality, depending on the genus. For instance, out of the 14 steps-awake duration variability interaction models, presence of 12 of the genera (which were mostly Clostridial) resulted in a modification of activity’s effect on awake duration variability, going from better to worse perceived sleep quality. Only *Eggerthella* and *Hungatella* modified the slope of steps on awake duration variability from positive (with microbe absent) to negative (with microbe present), improving the impact of steps on perceived sleep quality in participants with those microbes present. These taxa have previously been positively associated with depressive symptoms ^64^, but our findings show that independent of depression, presence of these taxa may benefit the host by modifying the effect of steps to decrease monthly awake duration variability, improving perceived sleep quality. This pattern of presence of Clostridial genera that modify the effect of activity effect to worsen perceived sleep quality contrasts with our main effects presence/absence and abundance model results that showed Clostridial genera are generally associated with better perceived sleep quality. For instance, across participants stratified by *Ruminococcus_1* presence/absence, participants who harbored *Ruminococcus_1* had overall better perceived sleep quality compared to participants without this genus, but within the *Ruminococcus_1* present group, the beneficial effect of activity on perceived sleep quality is diminished compared to those without the microbe. Another notable finding is that the presence of *Oscillospira* modified steps to decrease bedtime variability and therefore improve perceived sleep quality. *Oscillospira* also appeared in our main effects microbe abundance models where it was positively associated with sleep duration, consistent with another cross-sectional study ^68^, and in our presence/absence models where its presence was associated with decreased sleep disruption. These results suggest that *Oscillospira* not only supports longer and less disrupted sleep independent of activity level, but that it may also enhance the positive impact of activity on stabilizing bedtime, improving several aspects of sleep quality. *Oscillospira* has previously been proposed as a next-generation probiotic for supporting metabolic health ^69^, and our study is the first to highlight its potentially beneficial effects on sleep quality. *Lachnospiraceae_UC5_1_2E3*, on the other hand, was related to poor sleep quality in both the abundance and presence/absence models. This genus has previously been identified as a biomarker for worse metabolic outcomes, where increasing CLR-transformed abundance predicted higher values on a 2-hour insulin test ^70^. Sleep and metabolic health are closely intertwined ^71^, which is reflected by our findings of increased blood insulin and several blood lipids with worse perceived sleep quality. Taken together, our results suggest that microbes such as *Oscillospira* and *Lachnospiraceae_UC5_1_2E3* may influence sleep quality, within broader host metabolic pathways connecting the gut microbiome, sleep, and metabolic health.

Our study has several limitations. First, despite extensive covariate adjustments, the cross-sectional design of our analysis leaves open the possibility of residual confounding. For instance, we were not able to account for circadian timing of physical activity or eating. Second, reverse causality of sleep on molecular phenotypic data should be considered when interpreting our results. Sleep disruption may cause perturbations to gut microbiome or blood metabolome features, rather than vice versa. However, this does not preclude the possibility that findings presented here may still be useful therapeutic targets, given that sleep, physical activity, and the gut microbiome are known to be bidirectionally related to one another ^7,72,73^, and gut microbiome intervention trials have been shown to improve sleep quality ^73^. Third, our daily diet feature set analyses may have been underpowered due to small sample sizes (i.e., many individuals did not fill in their dietary questionnaires) and subject to recall bias, limiting our sensitivity to detect diet effects. Fourth, we did not include polynomial terms in our regressions, potentially missing non-linear associations. Finally, our cohort is a generally healthy, U.S.-based cohort, potentially limiting generalizability of findings to other populations. To address these limitations, future studies should consider collecting wearable-based longitudinal activity and sleep data with paired longitudinal diet and multi-omic data in diverse populations to work towards establishing directionality and causal mechanisms.

## Conclusion

Our findings demonstrate the utility of Fitbit devices to objectively measure sleep and activity at scale in free-living individuals in their natural settings. Monthly intra-individual average and standard deviation sleep metrics revealed dozens of associations with lifestyle and molecular phenotypic variables, independent of physical activity levels and genetic, demographic, and health status confounding. Reduced daily alcohol consumption and mineral supplement use appear to be practical interventions for improving sleep quality. Additionally, several butyrate-producing gut bacterial taxa, including *Coprococcus, Lachnospiraceae_UCG_001, Ruminococcaceae_UCG_003*, and *Christensenellaceae_R7_group*, emerged as potential targets to improve sleep quality in both depressed and non-depressed individuals, independent of their activity levels, and our results support emerging interest in developing next-generation *Parabacteroides* and *Oscillospira* probiotics ^69,74^. Conversely, the presence of *Lachnospiraceae_UC5_1_2E3* and its increasing abundance, along with higher fasting insulin levels, had a detrimental impact on sleep quality. Beyond main effects, we found that the presence and absence of certain gut microbes, including *Oscillospira*, moderated the effect of physical activity on sleep, identifying the gut microbiome as a potential target for precision probiotic interventions to modify how physical activity impacts sleep quality. Importantly, these associations were identified by our conservative analytical framework, where we required minimum sample size thresholds, extensive covariate adjustment, and false discovery rate control (q < 0.1). Our work provides targets for follow-up mechanistic studies and human intervention trials, and lays the foundation for personalized strategies to improve sleep quality.

## Methods

### Ethics and consent

Procedures for this study were reviewed and approved by the Western Institutional Review Board, with institutional review board study number 20170658 for the Institute for Systems Biology and 1178906 for Arivale.

### Arivale sample collection

Arivale participants were given Fitbit activity trackers and were administered diet, lifestyle, and clinical history questionnaires. Additionally, baseline blood draws were taken by trained phlebotomists at LabCorp or Quest service centres and paired with at-home stool sampling by Second Genome or DNA Genotek. Stool was processed for 16S amplicon sequencing (V4 region) by Second Genome or DNA Genotek. Blood plasma was processed for Olink proteomics and for global untargeted metabolomics by Metabolon.

### Fitbit data processing

The full Fitbit dataset showed five distinct missingness patterns that reflected heterogeneity in Fitbit APIs, device models, and software updates. Missingness patterns accounted for different variables (some participants did not have any recordings for certain variables over certain time periods) and different data distributions within the sleep_awakeningsCount variable. A categorical “missingness_pattern” variable was created to mark which missingness pattern each row belonged to in order to account for these Fitbit batch effects. Next, the full dataset was filtered based on information from the Fitbit Web API data dictionary (https://assets.ctfassets.net/0ltkef2fmze1/45IN5bvBS827grKEsA8ZB0/648f3778acc936961f057 2590c005ef0/Fitbit-Web-API-Data-Dictionary-Downloadable-Version-2.pdf and https://www.fitabase.com/media/2126/fitabase-fitbit-data-dictionary-as-of-05162025.pdf); the sleep_timeToFallAsleep variable should almost always be zero, so rows with non-zero values were dropped. The bedtime variable was derived from the sleep_startTime variable by shifting values from a 24-hour clock to a continuous 48-hour clock to preserve temporal ordering across midnight. For instance, bedtimes occurring between noon and midnight were retained as their original clock time (e.g., 13:30 was converted to 13.5), while bedtimes occurring after midnight were treated as occurring on the following day and were shifted by 24 hours (e.g. 1:45 was converted to 25.75). This conversion yielded a distribution of bedtimes ranging from 12 to 36, with an average bedtime of 23±2.2. Bedtime outliers, which were defined as outside 1.5*IQR, were dropped. Next, all “minutes” variables were summed to create a “total wear time” variable, and this variable was centered around 1440 (total minutes in a day). Total wear time outliers (outside of 1.5*IQR) were dropped.

After filtering, Fitbit data were windowed ±15 days from stool sample collection dates for each participant. Within this 30 day window, a weekend to weekday ratio was computed for each participant and their modal season was recorded. To ensure reliability of participant-level aggregates, a minimum of 7 Fitbit observations within the 30 day window for each participant was required to compute the average and standard deviation of a feature. The computed averages and standard deviations were then log(1+x) transformed and regressed on missingness pattern, modal season, the standard deviation of the Fitbit recordings’ ‘days a participant has been in the program,’ and weekend to weekday ratio. The residuals from these models were then saved to the working dataframe for all analyses.

### Data merging windows

Blood proteomics, untargeted blood metabolomics, and clinical blood chemistries were merged within ±15 days of a participant’s stool sample collection date. BMI and the digestion questionnaire were merged within 30 days, while the Oxford Happiness Questionnaire was merged within 50 days. The daily food frequency questionnaire, sleep disorder and mental illness questionnaires, self-reported supplements, Perceived Stress Scale scores (PSS), activity questionnaire, and sleep questionnaire were merged within 100 days of stool sample collection dates.

### Control covariate data in regression analyses

In addition to the sex, age, and BMI demographic covariates, the first three principal components of genetic ancestry were included in all models. These principal components were taken from a previous analysis of 100,000 linkage disequilibrium corrected SNPs (minor allele frequency >5%) calculated with the PC-AiR and PC-Relate methods ^75^. The history of a sleep disorder and history of a mental illness covariates were created by collapsing over several yes/no questionnaire items. The mental illness questionnaire assessed any current or previous diagnosis of bulimia, binge eating, anorexia, other eating disorder, anxiety, depression, general sadness, bipolar disorder, obsessive compulsive disorder, schizophrenia, other mental illness, attention deficit hyperactivity disorder (ADHD), alcohol abuse, and drug abuse. Importantly, questionnaire items differed substantially in terms of response completeness. The depression item had the highest total responses, and the depression, the anxiety, and the ADHD items had the highest “yes” counts and prevalences. Thus, the positive cases in the “history of a mental illness” covariate primarily consisted of individuals who reported depression, anxiety, and/or ADHD. The sleep disorder questionnaire had yes/no items asking about any current or previous diagnosis of insomnia, restless leg syndrome, sleep apnea, narcolepsy, or other sleep disorder. The same as the mental illness questionnaire items, there were substantial differences in the number of responses across sleep disorder items. The sleep apnea item had the most responses, and the sleep apnea, the insomnia, and the restless leg syndrome items had the highest “yes” counts and prevalences. Thus, the positive cases in the “history of a sleep disorder” covariate primarily consisted of individuals who reported sleep apnea, insomnia, and/or restless leg syndrome.

PSS scores ^76^ for each participant were treated as a continuous metric of psychological stress in all analyses.

Steps, floors, and activityCalories covariates measured the number of steps a participant took, the number of floors (elevation increase of 10 feet is considered one floor) climbed, and the number of calories burned beyond the user’s basal metabolic rate, respectively, in a day. More information on Fitbit features can be found on the Fitbit data dictionary (https://assets.ctfassets.net/0ltkef2fmze1/45IN5bvBS827grKEsA8ZB0/648f3778acc936961f057 2590c005ef0/Fitbit-Web-API-Data-Dictionary-Downloadable-Version-2.pdf and https://www.fitabase.com/media/2126/fitabase-fitbit-data-dictionary-as-of-05162025.pdf). The three monthly average Fitbit activity feature values were derived as described in the Fitbit data processing section above. The additional 17 Fitbit activity features and eight Fitbit heart rate features used in the random forest analysis (including both monthly averages and standard deviations) were derived using the same processing pipeline as the Fitbit activity and sleep features described above.

### Sleep, Activity, and Oxford Happiness Questionnaire data

The sleep questionnaire consisted of five items: average sleep, sleep hours, sleep trouble, sleep concentration, and restful sleep. Sleep trouble assessed how many nights per week were troubled sleep nights, while sleep concentration assessed how much a poor night’s sleep affected concentration the next day with responses ranging from “not at all” to “an extreme amount of impairment”. Restful sleep had response options of “yes”, “no”, and “unsure,” coded as 0, 1, and 2, respectively. The activity questionnaire set consisted of two items: moderate activity duration and vigorous activity duration. Each item asked participants to report how much time per day they spent engaged in moderate or vigorous physical activity. Both items had seven possible responses ranging in ascending order from “10 minutes” to “more than 60 minutes”, and an “N/A” option which was coded as 0 in the analysis. Sleep and activity questionnaire items were treated as independent variables predicting Fitbit sleep and activity features, respectively. Questionnaire items were modeled using an orthogonal polynomial coding scheme (https://www.statsmodels.org/dev/contrasts.html), in which an item with *k* ordered response levels was represented by k − 1 polynomial contrast terms (e.g., linear, quadratic, cubic). Only the linear term from each model was evaluated and reported. All models were adjusted for sex, age, BMI, the first three principal components of genetic ancestry, questionnaire vendor, and the time elapsed between Fitbit recordings and questionnaire completion. Sleep questionnaire items predicting Fitbit sleep features and activity questionnaire items predicting Fitbit activity features were treated as separate families of tests. Within each family, associations were further grouped by dependent Fitbit feature; for each Fitbit sleep (or activity) feature, FDR was controlled by applying the Benjamini-Hochberg procedure with the statsmodels multipletests function (https://www.statsmodels.org/dev/generated/statsmodels.stats.multitest.multipletests.html) to the set of p-values corresponding to all questionnaire items tested against that feature, with q-values less than 0.1 considered statistically significant ^77^.

Oxford Happiness Questionnaire responses were on an equally spaced Likert scale from 1 to 6, where 1 indicated “strongly disagree” and 6 indicated “strongly agree” with an item. Responses were converted to integers 1 through 6, and questionnaire items were treated as continuous dependent variables in all models. Models were adjusted for sex, age, BMI, and the first three principal components of genetic ancestry. For each questionnaire item, the FDR across the 18 sleep feature models was controlled to less than 10% by applying the Benjamini-Hochberg procedure to the set of p-values and retaining associations with q-values less than 0.1.

### Lifestyle and blood-derived multi-omics data

Food frequency questionnaire responses were converted to daily intake continuous variables by dividing the midpoint of the reported frequency by the time interval (e.g. “4-5 times per day” was converted to 4.5/1=4.5; “Once per week” was converted to 1/7=0.143). Digestion questionnaire items were left as-is. Self-reported supplements were identified from free-response boxes using regular expressions designed to detect supplement keywords and their spelling variants. Detection of a supplement was coded as 1, and failure to detect was coded as 0.

Data transformations, ln() or ln(1+x), were applied to all features from the untargeted blood metabolomics and clinical blood chemistries to reduce right-skew in variables.

### Stool 16S amplicon sequencing data

External providers performed amplification and library preparation, with 300-bp paired-end MiSeq profiling of the 16S V3 + V4 region (DNAgenotek) or 250-bp paired-end MiSeq profiling of the 16S V4 region (Second Genome). The FASTQ files were provided by the Illumina Basespace platform after the phiX reads were removed with basecalling. Length cutoffs of 250-bp for the forward reads and 230-bp for the reverse reads were employed. Further analysis for quality filtering, error modeling, and denoising was performed using the workflow from mbtools (https://github.com/Gibbons-Lab/mbtools) that wraps DADA2 ^78^. Taxonomy assignment was performed using the RDP classifier with the SILVA database (v132) ^79,80^. Here, 99% of the reads could be classified at the family level, 89% at the genus level (the taxonomic level chosen for this analysis), and 32% at the species level. This genus-level count table was used to compute center log ratio (CLR) transformed genus-level microbe abundance features, where zero counts were imputed with 0.5 prior to transformation ^81^. Imputed values were then replaced with “NaN” values post-CLR transformation. The “rrarefy” function in Vegan (v2.7-2) was used to generate a count table rarefied to 21,557 reads. Three diversity metrics were calculated as follows: (1) richness: number of observed genera for a given sample in the rarefied count table; (2) dominance (Berger-Parker): number of reads assigned to the most abundant genus in a sample divided by the total number of reads in that sample; and (3) Information (Shannon): H = 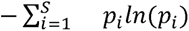, calculated using the “diversity” function in Vegan (v2.7.1). Presence/absence microbe features were also created from the rarefied count table, where “absence” was coded as 0 if a participant had a zero count for a particular microbe feature, or 1 for “present” if the count was greater than zero. Microbial community-scale metabolic models (MCMMs) were constructed for each participant in MICOM (v0.37.0) ^28^ using genome-scale metabolic models (GEMs) from the AGORA (v1.03) database summarized to the genus level ^82^. A relative abundance threshold of 0.001 was used, removing taxa below a relative abundance of 0.1% within a sample. MCMM growth was simulated using the cooperative_tradeoff() function in MICOM, applying a tradeoff parameter of 0.7. All simulations were run under a standard European diet, and metabolic production rates were computed using the micom.measures.production_rates() function. Metabolic production variables were natural logarithm-transformed, then saved as the MICOM metabolic output feature set and merged onto the working dataframe.

### Random forest variance partitioning analysis

For each predictor feature set (defined in Fig. S2), with the exception of the daily food frequency questionnaire set due to an insufficient number of complete samples, random forest regression models (max_features=0.33 and n_estimators=500) were fit to predict each monthly Fitbit sleep feature. In addition to the lifestyle and biological multi-omic feature sets, a control covariate predictor feature set was made of sex, age, BMI, the first three principal components of genetic ancestry, psychological stress (PSS), and history of a sleep disorder or a mental illness. The “All Features” set included all predictor feature sets combined. The Fitbit sleep features (defined in Fig. S2) were residualized on predictor feature set vendors, using linear regression, prior to random forest modeling. Model performance was evaluated using repeated K-fold cross-validation (four folds × 10 repeats). For each repeat, the dataset was randomly split into K=4 folds; models were trained on three folds and evaluated on the remaining fourth fold, such that each fold served as the test set once per repeat. This procedure was repeated 10 times, and the reported R^2^ values were the mean across all test folds of all repeats. The 95% intervals were defined as the 2.5th and 97.5th percentiles of the test fold R^2^ distribution. For each training set missing values in the predictor feature set variables were imputed with the median (or mode in the case of the digestion questionnaire feature set), and missing values in the testing sets were imputed with the training-derived imputed values. Analyses were performed in Python (v3.11.5) using the RandomForestRegressor and RepeatedKFold functions from the sklearn package (v1.3.1; https://scikit-learn.org/stable/supervised_learning.html). The “n_estimators” and “max_features” hyperparameters of the RandomForestRegressor function were set to 500 and ⅓, respectively, with the remaining hyperparameters set to their default settings.

### Main effects linear regression model specifications and workflow

Predictor features of interest (individual lifestyle and biological multi-omic factors) were organized into a total of 10 unique feature sets: daily diet questionnaire, self-reported supplements, digestion questionnaire, blood proteomics, untargeted blood metabolomics, clinical blood chemistries, gut microbiome alpha diversity, CLR transformed microbe abundances, microbe presence/absence, and MCMM-predicted metabolic output feature sets. There were 18 Fitbit sleep features, and for each predictor in a given feature set, all *Fitbit sleep ∼ predictor* combinations corresponding to the Cartesian product “Fitbit sleep feature set × feature set j” were fit as described in Figure S3, provided the specified model met all fitting criteria. For all models, at least N = 10 per independent variable was required to fit a regression model. In the case of binary predictors of interest (microbe presence/absence and supplement feature sets), the number of positive cases was required to be within 10-90% of the total N used to fit the model. For ordinal categorical predictors of interest (digestion questionnaire items), each response level had to have N ≥ 10 to fit a model.

Two models A and B were fit per *Fitbit sleep feature ∼ predictor* comparison. Model A regressions were specified as:

> *Fitbit sleep feature* ∼ *predictor* + sex + age + BMI + the first 3 principal components of genetic ancestry + the number of days between predictor sample collection and Fitbit recording + predictor vendor (where applicable).

Model B regressions were specified the same as Model A, with additional adjustment for psychological stress (PSS), history of a sleep disorder or a mental illness and the questionnaire vendor, and three Fitbit activity features (activityCalories, steps, and floors). Statistically significant predictors were identified by grouping predictor coefficient p- values by model (A or B), Fitbit sleep feature, and predictor feature set, and the Benjamini-Hochberg method was applied to each grouping to compute q-values from the p-values. For example, one test set for which q-values were computed was all A models which had minutes asleep as the outcome regressed on each digestion questionnaire feature from the digestion questionnaire feature set. Significant results in the model A set (q < 0.1) were then compared to significant results from the model B set (q < 0.1), and the overlapping significant results present in both models A and B were presented in the figures. The two models A and B differed in their extent of covariate adjustment, with the former including a baseline set of covariates, and the latter an expanded covariate set. We used this strategy to balance control for confounding with the potential for collider bias ^83^. Confounders are variables that are a common cause of a predictor (X) and an outcome (Y) ^83^. Failing to adjust for a confounder can result in a type I error, where the researcher wrongly concludes that X causes Y, but in reality X and Y are only related because they are both outcomes of a common cause ^83^. In contrast, collider variables are a common outcome of X and Y. Adjusting for a collider can cause a type I error by opening a non-causal pathway between X and Y ^83^. Because large covariate sets can inadvertently contain colliders, we chose to only retain significant associations that were present in both models A and B. Significant associations present in model B but not A may be spurious associations as a result of conditioning on a collider ^83^. For instance, the gut microbiome and poor sleep quality are linked to depression ^64^. If the “history of a mental illness” covariate lies on a pathway downstream of microbe features and Fitbit sleep metrics, including this variable in the regression models could introduce collider bias. All predictors were treated as continuous variables, except for the self-reported supplements, digestion questionnaire, and microbe presence/absence feature sets. The self-reported supplements and microbe presence/absence feature sets were treated as binary variables. Zero represented absence of a microbe or not taking a supplement, and 1 represented presence of a microbe or taking a supplement, with the reference group being 0. All digestion questionnaire items were treated as ordinal categorical variables and modeled using an orthogonal polynomial coding scheme (https://www.statsmodels.org/dev/contrasts.html), where an item with *k* ordered response levels was represented by *k - 1* polynomial contrast terms (linear, quadratic, cubic, etc.). The p-values and q-values for the digestion questionnaire items corresponded to the linear terms of these models. All analyses were conducted in Python (v3.11.5) using the ols function for model fitting and multipletests function for q-value calculations from the statsmodels package (v0.14.0; https://www.statsmodels.org/v0.14.0/user-guide.html).

### Interaction effects linear regression model specifications and workflow

All models examining interaction effects between Fitbit activity and microbe presence/absence were specified in the form:

> *Sleep feature ∼ activity feature : microbe presence/absence feature interaction term* + activity feature + microbe presence/absence feature + 16S sequencing vendor + sex + age + BMI + the first 3 principal components of genetic ancestry + the number of days between stool sample collection and Fitbit recording + history of a sleep disorder + history of a mental illness + mental illness and sleep disorder questionnaire vendor + microbiome vendor + psychological stress (PSS).

Models were fit for every combination of a Fitbit sleep feature, a Fitbit activity feature, and a microbe presence/absence feature, corresponding to the full Cartesian product “Fitbit sleep feature set × Fitbit activity feature set × microbe presence/absence feature set”. Two regression fitting criteria were enforced: a minimum 10 samples per independent variable in the model, and microbe presence between 10-90% of the dataframe used to fit the model.

Fitbit activity:microbe presence/absence interaction term p-values were grouped by sleep feature and activity feature, and the Benjamini-Hochberg procedure was applied to compute q-values. Only interaction terms with q-value less than 0.1 were considered statistically significant and presented in the figures. All analyses were conducted in Python (v3.11.5) using the ols function for model fitting and multipletests function for q-value calculations from the statsmodels package (v0.14.0).

## Data Availability

All ISB-generated and non-commercial datasets supporting the findings of this study are publicly available upon publication. These include microbiome sequencing data deposited in the NCBI Sequence Read Archive (SRA) under accession numbers PRJNA826530 and PRJNA826648, and metabolomics data deposited in MetaboLights under accession MTBLS2308. De-identified individual-level data derived from Arivale commercial subscribers are subject to legal and contractual restrictions under an Asset License Agreement between ISB and Arivale, Inc., and cannot be deposited in a public repository. Qualified researchers can access the full Arivale de-identified dataset supporting the findings in this study for research purposes through signing a data use agreement. Data access requests can be made at and will be responded to within 7 business days.

## Code Availability

Code and intermediate data files for reproducing the analyses and visualizations presented in this manuscript can be found in the following GitHub repository: https://github.com/Gibbons-Lab/2026_Cavon_lifestyle_and_multi-omic_determinants_of_sleep_quality

## Acknowledgments

We thank members of the Gibbons Lab for helpful discussions about this work. Figures were edited in https://BioRender.com. This work was supported by the National Institute of Diabetes and Digestive and Kidney Diseases (NIDDK) of the National Institutes of Health (NIH) under award number R01DK133468 and by a Global Grants for Gut Health Award from Nature Portfolio and Yakult (to SMG).

